# Alternative preservation strategies of DCD hearts: Effect of cold ischemia time before normothermic machine perfusion

**DOI:** 10.1101/2025.06.04.25329001

**Authors:** Manuela Lopera Higuita, Emmanuella O. Ajenu, Maya Bolger-Chen, George Olverson, Soheila Ali Akbari Ghavimi, Si Young Lee, Taku Kasai, William Michaud, Chukwudi Chijioke, Bhuban Ruidas, Allison Pitti, Nathan Minie, Joseph Catricala, Doug Vincent, Selena Li, David D’Alessandro, Elena Aikawa, Sasha A Singh, Marie Billaud, Asishana A. Osho, Shannon N. Tessier, S. Alireza Rabi

## Abstract

Donation after circulatory death (DCD) offers a promising strategy to expand the donor pool by 20–30%. However, DCD heart grafts suffer from warm ischemia injury, making effective graft viability assessment before transplantation prudent. Current methods for assessing DCD hearts are impacted either by ethical concerns and regional limitations (thoracoabdominal normothermic regional perfusion - taNRP) or by the need for significant resources, expertise, and equipment, with limitations in functional assessment (direct procurement and reperfusion – DPP, via the Organ Care System (OCS) by TransMedics). This study explores an alternative approach to DCD heart preservation, using controlled hypothermia at 6-8 degrees Celsius for up to 7 hours, followed by reanimation using normothermic machine perfusion. DCD grafts were procured from porcine donors after 19 mins of functional warm ischemia, stored via controlled cold static preservation for up to 7 hours and evaluated via a novel method of ex vivo cardiac loading. No functional, biochemical or proteomic differences were observed between DCD cardiac grafts stored via controlled hypothermia for up to 7 hours and DCD grafts reperfused immediately after procurement. These results suggest that controlled hypothermia may be a viable alternative to current DCD preservation methods, eliminating ethical concerns of taNRP and reducing logistical complexity and costs associated with DPP. These findings provide a foundation for a potential shift in DCD heart preservation practices, offering a more effective and cost-efficient approach to expand the donor pool and improve transplantation outcomes.

## Introduction

Heart failure is a chronic, progressive condition expected to affect over 8 million people in the United States alone by 2030 [1], with heart transplantation remaining the only definitive cure for end-stage heart failure [2]. However, broad access is significantly limited by the critical shortage of organ donors [3]. Various strategies have been explored to expand the donor pool, with donation after circulatory death (DCD) showing particular promise, as the utilization of DCD grafts has the potential to increase the organ donor pool by 20 – 30% [4]. DCD hearts undergo a mandatory period of warm ischemia (WIT) after the withdrawal of life support and during the donor’s progression to death. During this period, coronary perfusion pressure is insufficient to deliver adequate oxygen to the myocardium, leading to ischemia and possible dysfunction, necessitating assessment of graft viability prior to implantation.

Two methods are currently employed to assess DCD hearts post-WIT: thoracoabdominal normothermic regional perfusion (taNRP) and direct procurement and perfusion (DPP) [5]. Assessment via taNRP involves placing post-mortem donors on cardiopulmonary bypass while excluding cerebral circulation to avoid brain reperfusion [6]. After graft reanimation and a stabilization period, extracorporeal support is gradually removed, and the cardiac graft is evaluated using standard assessment tools [6]. Following evaluation, viable organs are subsequently transported using static cold storage (SCS), or normothermic machine perfusion (NMP) [7]. In addition to aiding in graft assessment, taNRP is suggested to improve post-transplant outcomes by mitigating the effects of warm ischemia, as ATP levels are restored and ischemic preconditioning may be activated [8]. Despite this advantage and being arguably more cost-effective than DPP [9], ethical concerns have hindered the widespread application of this assessment method, with some regions even imposing moratoriums [10, 11].

Alternatively, assessment via DPP involves rapid graft procurement following WIT, with subsequent reanimation on a commercially available NMP platform. The beating hearts are then transported by airplane to recipient centers and during this travel period, their viability is assessed based on lactate trends and visual inspection [12]. The immediate reperfusion via NMP limits cold ischemia and perfuses the heart with warm, oxygenated blood supplemented with nutrients [13, 14]. Furthermore, this technique enables some degree of ex vivo functional assessment through maintenance of the beating heart [15]. However, the implementation of DPP requires significant expertise, logistical coordination, and resources, which may be prohibitive for some transplant centers [13, 16]. Furthermore, although the Organ Care System (OCS) is the only FDA-approved NMP platform and is widely utilized, it lacks robust functional assessment capabilities. This is because the hearts are perfused in a non-working, unloaded, Langendorf mode, and its primary quantitative metric, lactate levels, has proven to be an unreliable predictor of heart function [17].

Despite its limitations, DPP remains the predominant method for DCD heart preservation and assessment in the United States and globally [18]. At the core of the DPP paradigm is the assumption that DCD hearts require immediate normothermic reperfusion to minimize further ischemic episodes and preserve graft viability [19]. However, the absolute necessity of immediate normothermic reperfusion remains unproven with limited direct evidence supporting the superiority of bypassing hypothermic preservation altogether [20–22]. On the contrary, emerging data suggest that a period of controlled hypothermia prior to NMP may improve graft function and reduce injury, as demonstrated in a rodent model of DCD heart transplantation, where grafts were exposed to oxygenated hypothermic perfusion following warm ischemia and prior to NMP [23]. Similarly, another study demonstrated that exposure to hypothermia attenuated the burst of free radicals and oxidative stress associated with reperfusion injury in rodent hearts [24].

The cardioprotective effects of hypothermia prior to reperfusion have also been well supported in the myocardial infarction literature. Decades of experimental and clinical evidence (reviewed in [25] and [26]) have shown that controlled hypothermia before reperfusion reduces reperfusion injury and dampens the inflammatory response, effectively preconditioning the myocardium for normothermic reperfusion. A similar trend has been observed in lung transplantation, where moderate hypothermic preservation (6–10°C) has been associated with improved graft function and post-transplant outcomes [27, 28].

The significant improvement in preservation technologies has enabled a more controlled hypothermic storage compared to the traditional SCS on ice. These emerging techniques allow us to maximize possible protective effects of hypothermia without the potential harmful effects of ice storage. As a result, transporting DCD hearts in static controlled hypothermia followed by NMP at the recipient center may offer distinct advantages over the current DPP strategy. This preservation method reduces the logistical complexity and cost associated with portable perfusion devices and in-transit NMP. Moreover, it enables more sophisticated functional assessments at the receiving transplant center, including cardiac loading protocols that offer a more comprehensive evaluation of graft viability prior to implantation [29].

To evaluate the plausibility of this preservation/assessment method, this study utilized a porcine model of donation after circulatory death, where grafts were procured and stored for up to 7 hours in controlled hypothermia using the Paragonix SherpaPak system. Following storage, grafts were reanimated via normothermic machine perfusion and maintained via a novel loaded perfusion mode which simulates near-physiologic conditions [29]. Grafts were then evaluated via a comprehensive array of biochemical, proteomic, imaging, and functional tests. This study sets a precedent for potentially shifting the current DCD heart preservation paradigm away from either taNRP and DPP and towards a more effective and less costly approach.

## Methods

All research complies with the ethical standards and regulations set forth by National Research Council guidelines and in the experimental protocol 2023N000042 approved by the Institutional Animal Care and Use Committee (IACUC) of Massachusetts General Hospital (Boston, MA, USA) and the ARRIVE guidelines.

### Porcine Model of donation after circulatory death

Yorkshire pigs (30–35 kg, age 3–4 months, mixed sex) were anesthetized and intubated, with anesthesia maintained using inhaled isoflurane (3%–5%) and intravenous fentanyl (5–20 µg/kg/h) as needed. After the full depth of anesthesia was confirmed, midline sternotomy was performed. Systemic heparin (300 U/kg) was administered and allowed to circulate for 3 minutes. A cardioplegia needle was placed in the ascending aorta. A pressure sensor was connected to a side channel of the cardioplegia cannula and used to continuously monitor the mean arterial pressure (MAP). Once proper pressure monitoring was confirmed, the animals were transitioned to intravenous anesthesia, ventilation turned off and the animals were allowed to expire secondary to apnea. Once the MAP dropped below 30 mmHg, functional warm ischemia time (fWIT) was initiated and observed for a total of 19 mins, during which asystole occurs. After 19 mins of fWIT, 1L of blood is collected from the right atrium to be used during normothermic perfusion. The left and right atria were vented, and cross clamp was placed distal to the cardioplegia cannula, the heart was flushed with 1L of del Nido cardioplegia solution at 4°C. Hearts preserved via controlled hypothermic storage before NMP were then flushed with an additional liter of University of Wisconsin (UW) solution, also at 4°C. Following the flush, cardiotomy was performed in a standard fashion.

Hearts were either immediately instrumented for placement on the normothermic machine perfusion system (Minimal cold ischemia time – CIT, n = 6) or stored using controlled static hypothermic preservation in 3-4L of UW solution within the Paragonix SherpaPak (Waltham, MA) for 5 hours (5 h controlled cold storage preservation – cCSP, n = 7) or 7 hours cCSP (n = 6, Figure 1). Hearts stored under cold ischemia conditions were subsequently instrumented for placement on the ex vivo perfusion platform.

**Figure 1:**
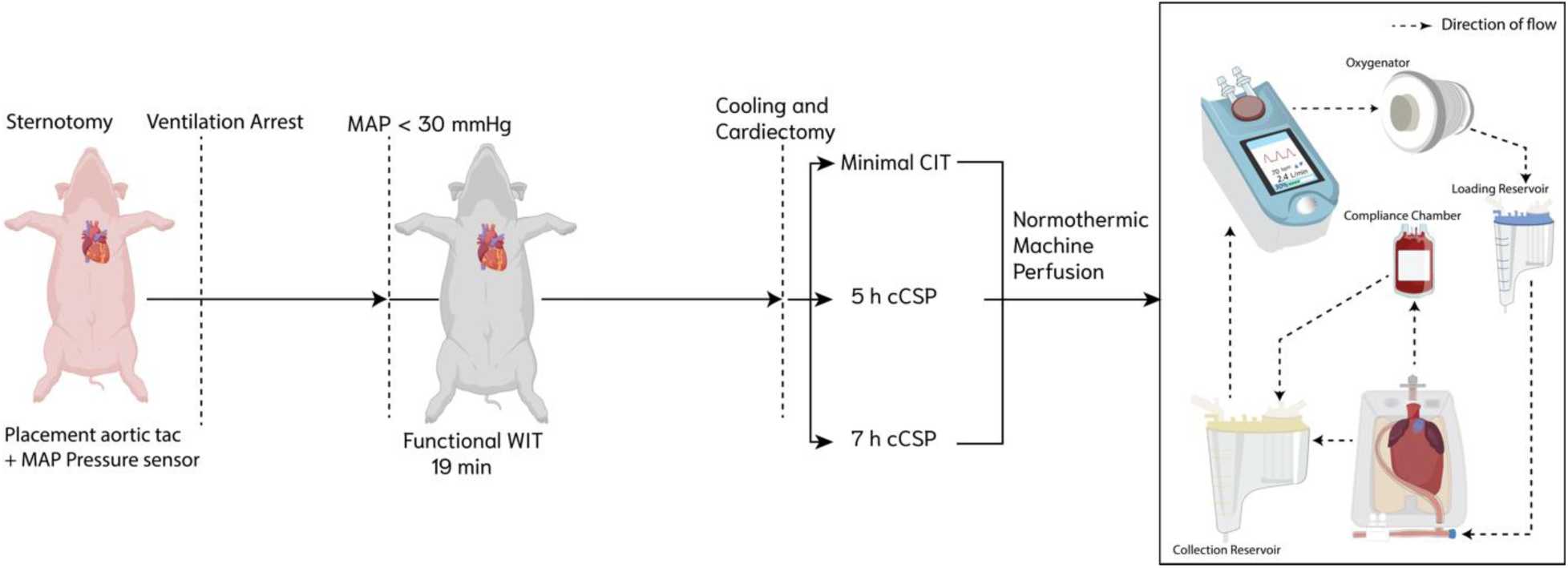
Experimental setup and design. Donation after circulatory death was imitated by allowing pigs to expire due apnea after ventilatory arrest until a mean arterial pressure (MAP) of 30 mmHg was reached. Once the target MAP was achieved, functional warm ischemia time was maintained for a total of 19 mins. Hearts were either reperfused immediately via normothermic machine perfusion for evaluation or stored via controlled cold storage preservation (cCSP) for 5 hours or 7 hours. Once storage time was completed cCSP hearts were reperfused via normothermic machine perfusion for evaluation.

### Heart Instrumentation

The left atrial (LA) vent site was sutured closed with 4-0 prolene continuous suture, pacing wires were attached to the myocardium, and a double purse string suture was placed around the LA cuff to allow sealing and achieve LA pressurization during loaded mode perfusion. The aorta was cut below the arch, and an aortic cannula (VentriFlo Inc, Pelham, NH) was inserted and fastened with a zip tie. The hearts were weighted and full thickness, 5 mm biopsy punches were obtained from the right ventricle for downstream analysis.

### Ex vivo perfusion platform

The ex vivo perfusion system, as well as the Langendorf and Loaded perfusion methods utilized in this study have been described in detail elsewhere [29]. Briefly, the system is comprised of an organ chamber (VentriFlo Inc, Pelham, NH) connected to a reservoir, which is, in turn, connected to a pediatric oxygenator (Terumo Medical Corporation, Somerset, NJ) via a pulsatile pump (VentriFlo Inc, Pelham, NH). During Langendorf perfusion, the oxygenator supplies with oxygenated perfusate a Windkessel bag (VentriFlo Inc, Pelham, NH) which is connected directly to the aorta. During Loaded perfusion, the oxygenator fills a secondary reservoir (Loading reservoir) which is utilized to pressurize the LA.

### Heart Perfusion

All chemicals were purchased from Sigma Aldrich unless otherwise stated. All hearts were retrogradely perfused for 1 hour, followed by 3 hours in Loaded mode in the aforementioned system. Hearts were perfused with 2 L of base perfusate containing 0.96% Krebs-Henseleit buffer, 9.915mM dextran, 25mM sodium bicarbonate, 1.054mM bovine serum albumin, 1% Penicillin-Streptomycin (10,00U/mL), 0.13% insulin (100U/mL), 0.02% hydrocortisone (50mg/mL), 0.5% heparin (1000 U/mL), and 2.75mM calcium chloride and 1 L of isolated red blood cells (1:2), warmed at 37°C by the circulation of warm water (Adroit Medical Systems, Loudon, TN) through the double jacket of the oxygenator and oxygenated with 100% O_2_. Isolated red blood cells were obtained via cell saver processing (Medtronic, Dublin, Ireland) of the whole blood collected during surgery, followed by filtration through a leukoreduction filter (Haemonetics, Boston, MA). Hearts were continuously administered adenosine (333 ug/min) delivered via a syringe pump connected to the system as close as possible to the organ.

After instrumentation, hearts were connected to the perfusion system maintaining an aortic pressure between 35 and 40 mmHg. Hearts were rhythmically massaged to avoid left ventricular distention until intrinsic contractions were detected. In case of fibrillation, hearts were defibrillated (Hewlett Packard, Louisville, KY) starting with 10 J, with subsequent shocks administered in increments of 10 J until rhythmicity was observed. A back up pacer was set at 60 beats per minute. Once stable conditions were achieved, three EKG leads were placed on the surface of the hearts, and the resulting signal transferred to the pulsatile pump to achieve counter pulsation between the pump and the heart. Hearts were perfused in retrograde mode for 1 hour.

### Loaded Mode perfusion

To achieve Loaded Mode, the Loading reservoir was filled until a pressure between 12 and 14 mmHg was achieved. The Loading pressure was maintained solely by controlling the fluid height in the Loading reservoir, leading to passive left atrial pressurization (i.e., gravity-dependent pressurization) [29]. A right-angle metal tip cannula (Medtronic) was inserted into the LA with the opening pointing towards the appendage to avoid obstruction of the mitral valve. Along with the canula, a fiber optic pressure sensor (304µm OD, Opsens, Canada) was inserted through the opening of the LA cuff and into the left ventricle for LV pressure recording. The LA cuff was closed around the canula and pressure sensor to create a seal by tightening the previously placed purse string sutures through snares. In the Loaded Mode perfusion implemented in this study, pump outflow is completely redirected towards the loading reservoir resulting in a passive afterload (i.e., perfusion of the coronaries and the mean arterial pressure is solely dependent on LV ejection without mechanical pump assistance) [29].

### Viability Assessment

Oxygen and lactate measurements were quantified from perfusate samples via a blood gas analyzer (Siemens Medical Solutions, Malvern, PA, USA) every 30 mins. Oxygen uptake rate (OUR) was determined as described previously [30]. Hearts were weighed before and after perfusion, and these values were utilized to calculate percent weight gain, a proxy for edema.

Mitochondria redox state during perfusion was quantified via Raman resonance spectroscopy (Pendar Technologies, Cambridge, MA), as extensively described elsewhere [31]. Briefly, the device uses a 441 nm laser to measure Raman resonance photons from the left ventricle, with data collected over 180 seconds. A custom LabView software subtracts the fluorescence baseline and applies a regression algorithm to demultiplex the spectrum, using a library of chromophore spectra (mito-reduced, mito-oxidized, and myoglobin) to analyze the data. The regression calculates the relative concentrations of metabolic molecules, allowing the determination of the 3RMR ratio, which represents the proportion of reduced mitochondria to total mitochondria.

The energetic status of cardiac cells was calculated via the energy charge of the adenylate system following the formula: (ATP + 0.5*ADP)/(ATP + ADP + AMP) [32]. The metabolic cofactors (ATP, AMP, and ADP) were obtained via mass spectrometry, as described previously [33]. Briefly, biopsy punches obtained from the right ventricle before and after perfusion were flash-frozen in liquid nitrogen, and stored at -80°C. Flash-frozen tissue was homogenized and analyzed using a 3200 triple quadrupole liquid chromatography-mass spectrometry system (AB Sciex, Toronto, Canada).

Mitochondria morphology was assessed from electron microscopy images. Pre- and post-perfusion samples (1mm x 1mm) were fixed for several hours at room temperature in 2.0% paraformaldehyde/2.5% glutaraldehyde in 0.1M sodium cacodylate buffer (pH 7.4), then incubated in fresh fixative overnight at 4°C. Specimens were rinsed several times in 0.1M cacodylate buffer, incubated for 1hr in 1% osmium tetroxide, rinsed again multiple times in cacodylate buffer, then dehydrated through a graded series of ethanol incubations until reaching 100%, followed by a 10mins dehydration in 100% propylene oxide. Tissue was then transferred into a 2:1 mix of propylene oxide:Eponate resin (Ted Pella, Redding, CA), incubated for 2 hours at room temperature, then placed into a 1:1 mix of propylene oxide:Eponate resin, incubated overnight at room temperature on a gentle rotator. The following day, specimens were transferred into fresh 100% Eponate and incubated for several hours on a gentle rocker table, then embedded in flat molds with fresh 100% Eponate resin and allowed to polymerize in a 60°C oven (24-48hrs). Ultra-thin (70nm) sections were cut using a Leica EM UC7 ultramicrotome, collected onto formvar-coated grids, stained with 2% uranyl acetate and Reynold’s lead citrate and examined in a JEOL JEM 1011 transmission electron microscope at 80 kV. Once processed, 10-15 images per sample (n = 3 per CIT) were acquired using an Tecnai G2 Spirit BioTWIN transmission electron microscope (FEI, Hillsboro, OR) equipped with a Nanosprint 43-MKII camera (Advanced Microscopy Techniques, Woburn, MA). The mitochondria on each image were counted and their length and width quantified via ImageJ [34]. Average mitochondrial counts were manually scored in randomly selected fields. Mitochondrial dimensions including length and width were selectively measured, and viability was assessed based on intact membranes and clearly defined cristae.

Histological images were obtained from before and after punch biopsies fixed in 4% PFA (ThermoFisher, Waltham, MA) in PBS for one hour, rinsed twice with PBS, transferred to 70% ethanol, and embedded in paraffin. Embedded samples were cut and stained with hematoxylin and eosin stain.

### Functional assessment

Vascular resistance was calculated by dividing the coronary pressure by coronary flow rate and corrected for pre-perfusion graft weight. Left ventricular output pressure was acquired from the fiber optic pressure sensor inserted into the left ventricle during Loaded Mode perfusion and continuously recorded using Labscribe software (iWorx, Dover, NH, USA), and the data processed with MATLAB (MathWorks Inc, Natick, MA). Heart rate was calculated by counting the number of peaks per second from the left ventricular pressure recordings and multiplied by 60 to obtain beats per minute. Atrial and aortic pressures and flows were recorded every 30 mins and utilized to calculate the Loadability metric.

### Pro-inflammatory assessment

Pro-inflammatory markers were measured via bead-based immunoassay (Luminex, Austin, TX), by Eve Technologies Corp (Alberta, CA). Briefly, inflow perfusate samples were collected at 30 mins (T0.5) and 4 hours (T4), centrifuged at 2200 rpm for 10 mins to separate the red blood cells, and the acellular perfusate was flash frozen and shipped on ice for processing. Analysis was performed using a Luminex 200 system, following the kit’s manufacturer instructions (MilliporeSigma, Burlington, MA).

### Proteolysis

Proteins lysates were prepared as described below on myocardium from the left and right ventricles (LV, RV, respectively) harvested before and after NMP in hearts reperfused immediately after harvesting (Minimal CIT) and after storage and after NMP for hearts stored in cold conditions for 5 or 7h (cCSP). A total of 44 samples were processed. Protein extraction was performed on ice using a lysis buffer consisting of RIPA buffer (Thermo Scientific, Cat #89900), phenylmethylsulfonyl fluoride (PMSF, 1 mM, MP Biomedicals), protease inhibitor cocktail (Sigma Aldrich), and phosphatase inhibitor cocktail (Thermo Scientific). 44 myocardial specimens from the left and right ventricles (LV, RV) were cut into 2–4 mm pieces, mixed with lysis buffer (1 mL for 300 mg of tissue) and mechanically dissociated using the GentleMACS Dissociator (Protein01 program, Miltenyi Biotech). After centrifugation (100 g, 3 mins, 4°C), the tissue lysates were transferred to microcentrifuge tubes and underwent five freeze-thaw cycles. Lysates were further centrifuged (500 g, 5 mins, 4 °C) and a fraction of the supernatant was stored for future analyses while the other was re-centrifuged (21,300 g, 20 mins, 4°C). The protein concentration of the resulting fraction was determined using a bicinchoninic acid (BCA) assay (Thermo Fisher Scientific) according to the manufacturer’s instructions. Protein lysates were stored at −80 °C until peptide preparation. The lysates were proteolyzed using the iST in-solution digestion kit (PreOmics GmbH, Germany) automated on the PreON robot (PreOmics): 10 µg protein in 10 µL was added to 40 µL LYSE buffer (PreOmics). The samples were trypsinized for 3 hours following the manufacturer’s instructions. Eluted peptides were dried in a speed vacuum (EppendorfVacufuge) and resuspended in 40 µL LC-load.

### Mass spectrometry

Mass spectra were acquired on Orbitrap Fusion Lumos coupled to an Easy-nLC1000 HPLC pump (Thermo Fisher Scientific). The peptides from 44 samples were diluted 5-fold using LC-load and 4 uL injections separated using a dual column set-up: an Acclaim™ PepMap™ 100 C18 HPLC Column, 75 µm X 70 mm (Thermo Fisher Scientific, Cat# 164946); and an EASY-Spray™ HPLC Column, 75 µm X 250 mm (Thermo Fisher Scientific, Cat# ES902). The column was heated at a constant temperature of 45 °C. The gradient flow rate was 300 nL/min from 5 to 21% solvent B (0.1% formic acid in acetonitrile) for 75 minutes, 21 to 30% solvent B for 15 mins, and another 10 mins of a 95%-5% solvent B sawtooth wash. Solvent A was 0.1% formic acid in mass spectrometry-grade water. The mass spectrometer was set to 120,000 resolution, and the top N precursor ions in a 3 second cycle time (within a scan range of m/z 400-1500; isolation window, 1.6 m/z) were subjected to collision-induced dissociation (CID, collision energy 30%) for peptide sequencing.

### Mass spectral analyses

The acquired peptide spectra corresponding to the 44 samples were searched with Proteome Discoverer package (PD, Version 2.5) using the SEQUEST-HT search algorithm against the Sus Scrofa (Pig) Uniprot fasta database (downloaded March 19, 2024; n=22,784 entries). The digestion enzyme was set to trypsin and up to two missed cleavages were allowed. The precursor tolerance was set to 10 ppm and the fragment tolerance window to 0.6 Da. Methionine oxidation and n-terminal acetylation were set as dynamic modifications, and cysteine carbamidomethylation as a static modification. The peptide false discovery rate (FDR) was calculated by the PD Percolator algorithm and peptides were filtered based on an FDR threshold of 1.0%. Unique peptides were used to assign the Master protein and razor peptides were included for quantification. A minimum of 2 unique peptides for each protein were required for the protein to be included in the analyses. The Feature Mapper was enabled in PD to quantify peptide precursors detected in the MS1 but may not have been sequenced in all samples. Chromatographic alignment was performed with a maximum retention time shift of 10 mins, mass tolerance of 10 ppm and signal-to-noise minimum of 5. Precursor peptide abundances were based on their chromatographic intensities and total peptide amount was used for PD normalization. After exporting the data from Proteome Discoverer, a median normalization step was applied using a custom script in Python to fill missing quantification values with ‘0’ and rescale the data.

### Statistical Analysis

All data were analyzed for outliers using the ROUT method with Q = 1%, statistical outliers were excluded from data graphs. All results utilized 6 hearts in minimal CIT, 7 hearts in 5 hours, and 6 hearts in 7 hours groups unless otherwise indicated in the results section. Over-time data were analyzed using repeated measures two-way ANOVA and Tukey-Kramer HSD post-hoc analysis on standard least squares means. Column data were analyzed via Wilcoxon/Kruskal-Wallis Test with Dunn post-hoc analysis on non-parametric medians. Inflammatory marker data was analyzed via Difference in Difference test. To do so, it was assumed that in the absence of treatment with cCSP, the two groups of hearts will have similar changes in the inflammatory cytokine production during NMP. Therefore, any differences seen in the cytokine accumulation during NMP is attributed to one group being subject to cCSP. Statistical significance is defined as p< 0.05. For the purpose of robust power, we combined all the hearts preserved with cCSP under one group and compared them to the minimal CIT group. All data are expressed as median ± interquartile range (IQR). Statistical significance is defined at p < 0.05.

Perseus v 2.1.3 was used for group comparison of protein abundance after log2-transformation [35]. The quality of the dataset was ensured by plotting the frequency distribution of each protein abundance into a histogram for each sample. The Student’s t-test was used to calculate significantly different proteins between two groups (minimal CIT vs. CSP and post-storage vs. post-perfusion) at a permutation-based false discovery rate (FDR or adjusted q-value) ≤ 0.05. Volcano plots were generated using the results of the Student’s t-test. Principal component analyses were performed on complete proteomes while hierarchical clustering was performed on proteins found to be significantly different by Student’s t-test.

Gene ontology analysis was performed in DAVID https://davidbioinformatics.nih.gov/summary.jsp on proteins found to be significantly differentially expressed between groups [36]. Gene set enrichment analysis (GSEA) was performed using GSEA v4.4.0 and MSigDB indicated in the supplemental tables (S7-S10) [37, 38].

## Results

### Controlled hypothermic preservation of DCD grafts prior to NMP does not affect graft metabolism or function

Immediate normothermic machine perfusion is the bedrock of the DPP method of preserving DCD hearts, with the assumption that any period of cold ischemia between procurement and normothermic reperfusion is deleterious to the graft function [19]. However, the results in this study, along with a few other recent publications [20–22], challenge this assumption by demonstrating the function of DCD cardiac grafts is retained after hypothermic storage (Fig. 2 & 3).

**Figure 2:**
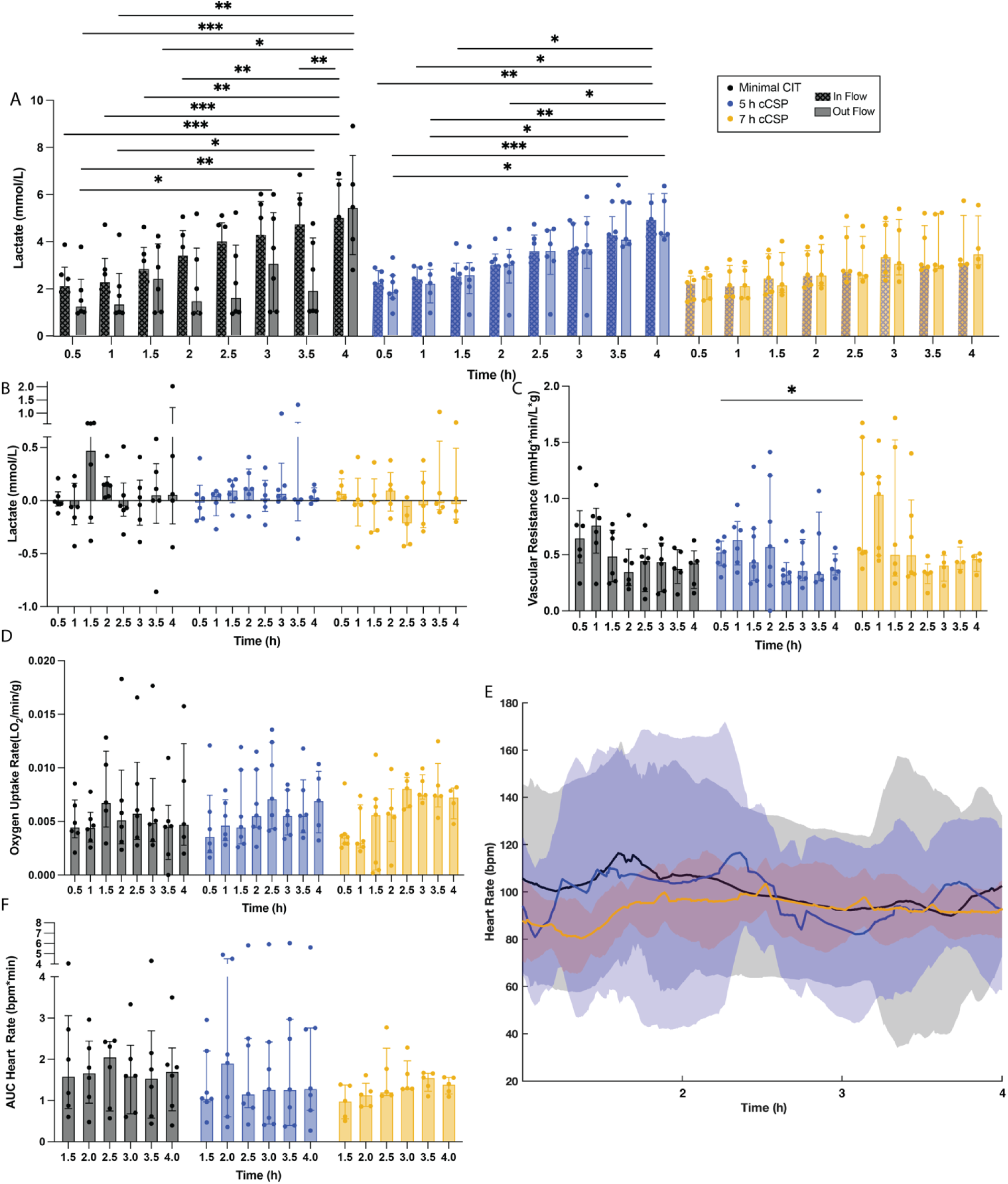
Conventional assessment metrics of graft viability. Metrics are considered conventional if their acquisition is possible during unloaded (e.g. Organ Care System) perfusion. (A) In perfusate InFlow (checkered columns) and OutFlow lactate (solid columns) measured overtime for Minimal CIT, 5 hours controlled cold storage preservation (cCSP) and 7 hours cCSP. (C) Coronary vascular resistance. (D) Oxygen uptake rate. (E) Heart rate, obtained from intraventricular pressure recordings. Solid line indicates the median of the experimental replicates, the shaded area indicates interquartile range. (F) area under the curve (AUC) of heart rate data for every half an hour of perfusion. Colum data presented as median ± interquartile range. Wilcoxon/Kruskal-Wallis Test with Dunn post-hoc analysis on non-parametric medians. Repeated measures two-way ANOVA and Tukey-Kramer HSD post-hoc analysis on standard least squares means. * = p <0.05, ** = p ≤ 0.01, *** = p ≤ 0.001.

**Figure 3:**
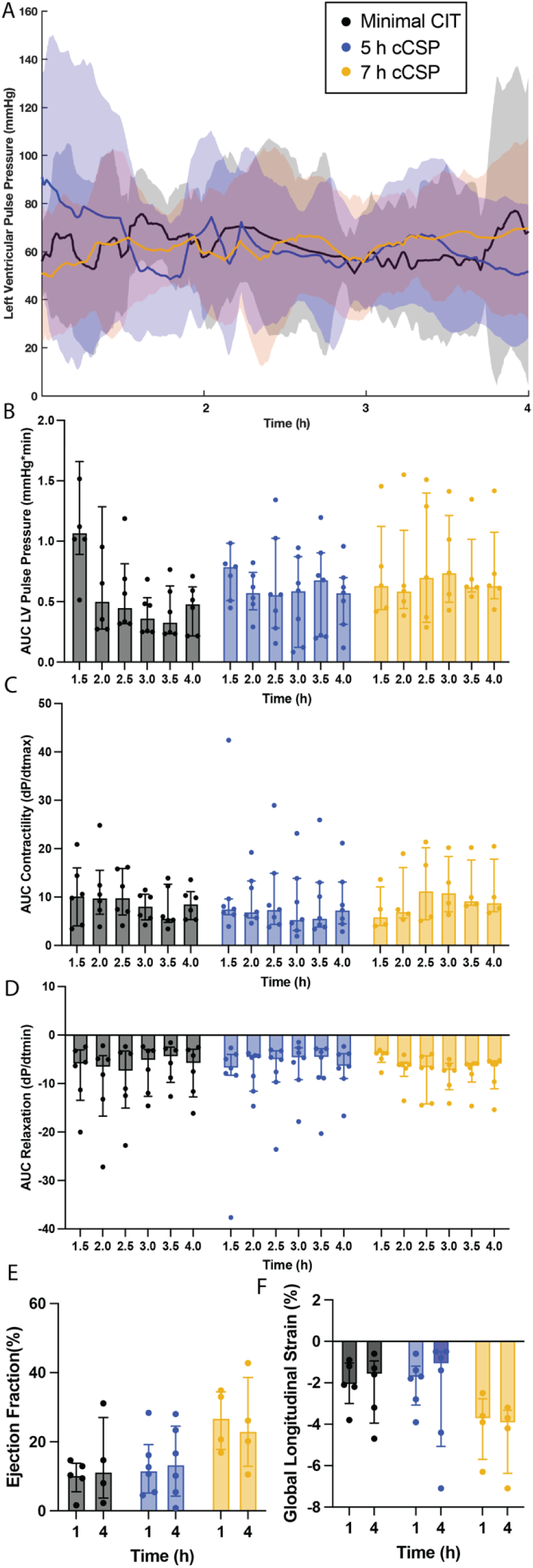
Nonconventional assessment metrics – left ventricular pulse pressure. Metrics are considered non-conventional if their acquisition is only possible via left ventricular loading and not common practice clinically. (A) Maximum systolic pressure over time, denoted as left ventricular pulse pressure (LVPP) of Minimal CIT (1 h CIT), 5 h controlled cold storage preservation (cCSP) and 7 hours cCSP. Solid line indicates median of the experimental replicates, the shaded area indicates interquartile range. (B) Area under the LVPP curve (AUC) for every half an hour of perfusion, (C) Cardiac muscle contractility quantified from the maximum derivative of the pressure pulse. (D) Cardiac muscle relaxation quantified from the minimum derivative of the pressure pulse. (E) Ejection fraction and (F) Global longitudinal strain obtained from epicardial echocardiographic. Colum data presented as median ± interquartile range. Wilcoxon/Kruskal-Wallis Test with Dunn post-hoc analysis on non-parametric medians. Repeated measures two-way ANOVA and Tukey-Kramer HSD post-hoc analysis on standard least squares means.

Both conventional (e.g., available during unloaded perfusion, as in the OCS platform) and non-conventional (e.g. functional assessment requiring left ventricular loading) metrics were used to assess graft metabolic fitness and function. An overall increase in lactate was seen in grafts in all groups (Fig. 2A), with the final concentration of lactate (T4) in Minimal CIT and 5ours cCSP grafts being statistically higher than initial lactate levels (T0.5). Alternatively, although still experiencing a progressive increase in lactate over time, the final lactate levels (T4) of 7 hours cCSP grafts were not statistically higher than levels at the beginning of perfusion (T0.5). Lactate consumption (outflow – InFlow, Fig. 2B) and vascular resistance (Fig. 2C) demonstrated no significant difference between groups of within groups over time. Similarly, no changes over time were observed in the oxygen uptake rate (OUR) or between groups (Fig. 2D). Likewise, no statistically significant differences in heart rate (Fig. 2E & 2F) were observed either over the course of perfusion or between groups.

Despite the slight variability in conventional assessment metrics, non-conventional, functional metrics of graft viability were comparable across all groups. No statistical differences were seen in left ventricular pulse pressure (systolic – diastolic, Fig. 3A & 3B), cardiomyocyte contractility (Fig. 3C) or relaxation (Fig. 3D) over time of between groups. Although ejection fraction was significantly lower (∼20%) than normal (50%–70%) [39], likely due to the non-physiological characteristics of the perfusion system, no differences in ejection fraction (Fig. 3E) or global longitudinal strain (Fig. 3F), obtained via epicardial echocardiography, were seen across groups immediately after left ventricular loading and after 4 hours of perfusion.

### Moderate hypothermic preservation prior to NMP does not adversely affect myocardial structure or ultrastructure

In addition to biochemical and functional assessments, myocardial structural integrity can serve as a marker of preservation quality. Extended cold ischemia on ice has been associated with cellular and ultrastructural damage [40]. No structural abnormalities were observed in hearts preserved at 6–8°C for 5 or 7 hours prior to NMP, as determined in H&E stained tissue, with visibly normal cardiomyocyte size, regular striations, uniform nuclei and normal interstitial spacing (no storage-induced edema) present in the right ventricle regardless of cold ischemia time (Fig. 4A). Slight disruptions are present after NMP both in the right (Fig.4B) and the left (Fig. 4C) ventricle of the cardiac grafts in all groups, with visibly increased irregularities and variability in cardiomyocyte orientation, cytoplasmic vacuolation, sporadic disruptions to striations, as well as the presence of red blood cells in the interstitial spaces, potentially indicating vascular congestion. Additionally, widening of the inter-myocyte spaces translated into weight gain, a proxy for edema, at the end of the 4 hours of perfusion (Fig. 4D). Minimal CIT grafts experienced a 33.81 % [10 – 46.67] weight increase, 5 hours cCSP grafts showed a 45.91 % [19.9 – 56.8] weight increase, and 7 hours cCSP grafts exhibited a 33.37 % [19.9 – 47.62] weight increase, with no statistical difference between the groups regardless of cold ischemia time (p = 0.214, Fig. 4D). The lack of differences in histological results across groups, both post-storage and post-perfusion, as well as in the quantified post-perfusion edema indicates these adverse changes are a direct effect of the NMP.

**Figure 4:**
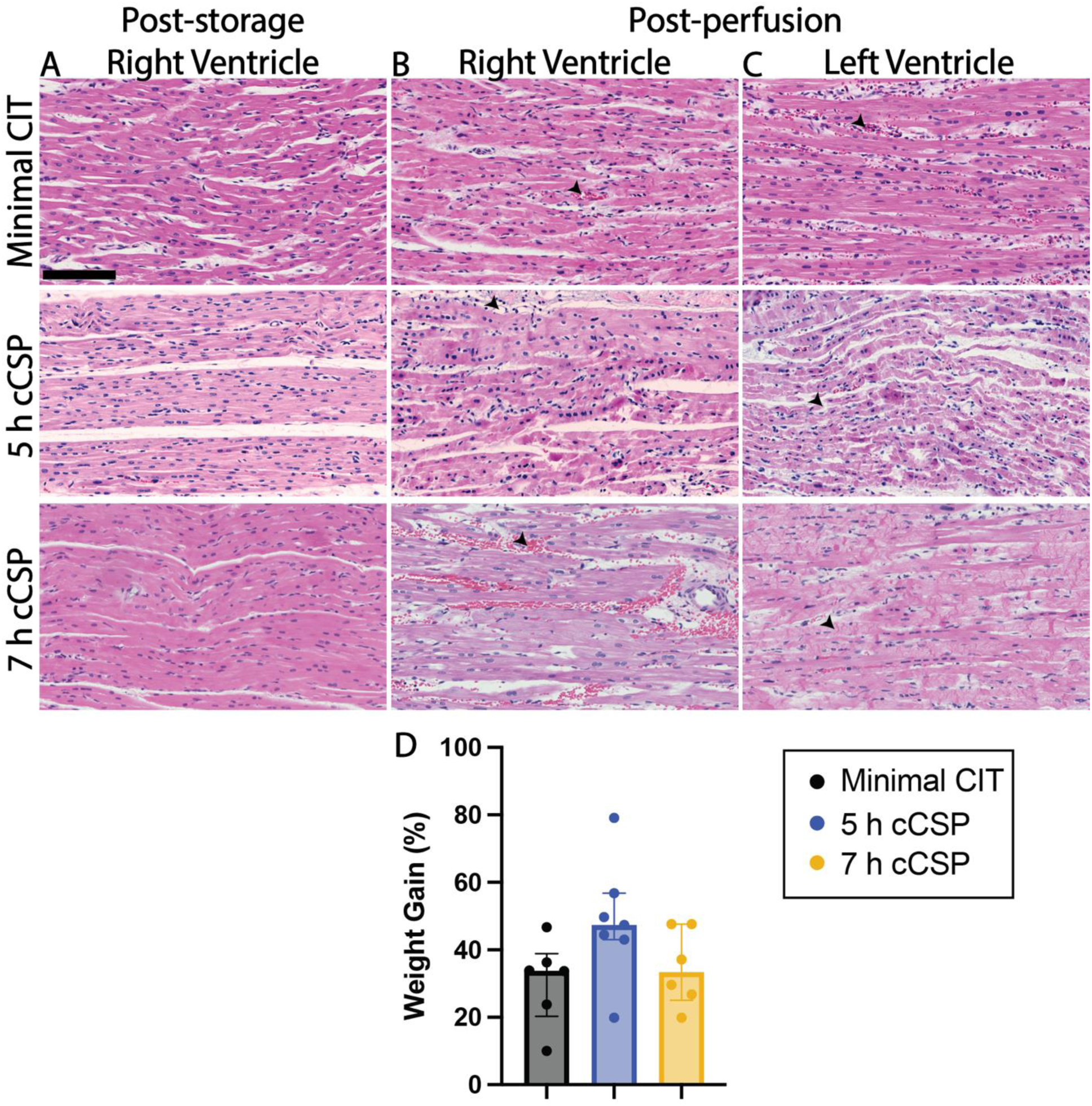
Effects of cold ischemia storage and NMP in myocardial structure and edema. (A) Representative H&E images of the right ventricular wall post-storage. Representative H&E images of the right (B) and left (C) ventricular walls post-perfusion. Scale bar 100 µm. (D) percent weight gain after 4 hours of perfusion. Colum data presented as median ± interquartile range. Wilcoxon/Kruskal-Wallis Test with Dunn post-hoc analysis on non-parametric medians.

### Mitochondrial ultrastructure, redox state and myocardial energy stores are preserved following moderate hypothermic preservation

Mitochondria are central regulators of myocardial injury during ischemia and reperfusion, with their health and structure directly influencing the extent of tissue damage [41, 42]. No mitochondrial ultrastructural abnormalities were observed in transmission electron microscopy images in hearts preserved at 6–8°C for 5 or 7 hours compared to those reperfused immediately (Minimal CIT) with regular size/shape, densely packed and organized cristae and intact mitochondria membranes (Fig. 5A). Similarly, no ultrastructural changes were seen after 4 hours of perfusion (Fig. 5B). Mitochondrial morphology including length (Fig, 5C) and width (Fig. 5D), as well as the number of viable mitochondria (Fig. 5E) remained unchanged, with no significant differences between groups in either post-storage or post-perfusion samples. The retention of mitochondria ultrastructure, shape and viability resulted in retained mitochondrial function, with no statistical differences between groups in mitochondria redox state (Fig. 5F), although statistically more reduced mitochondria (higher 3RMR) were seen in Minimal CIT grafts at 3 hours of perfusion when compared to earlier timepoints. Similarly, no significant difference was observed in energy charge between groups at either timepoint (Fig. 5G), with pre-perfusion and post-perfusion RV samples showing comparable energy content irrespective of cold ischemia time. These findings indicate that moderate hypothermic preservation does not impair mitochondrial function or reduce myocardial energy reserves, supporting the overall structural, ultrastructural, and metabolic integrity of DCD hearts prior to NMP.

**Figure 5:**
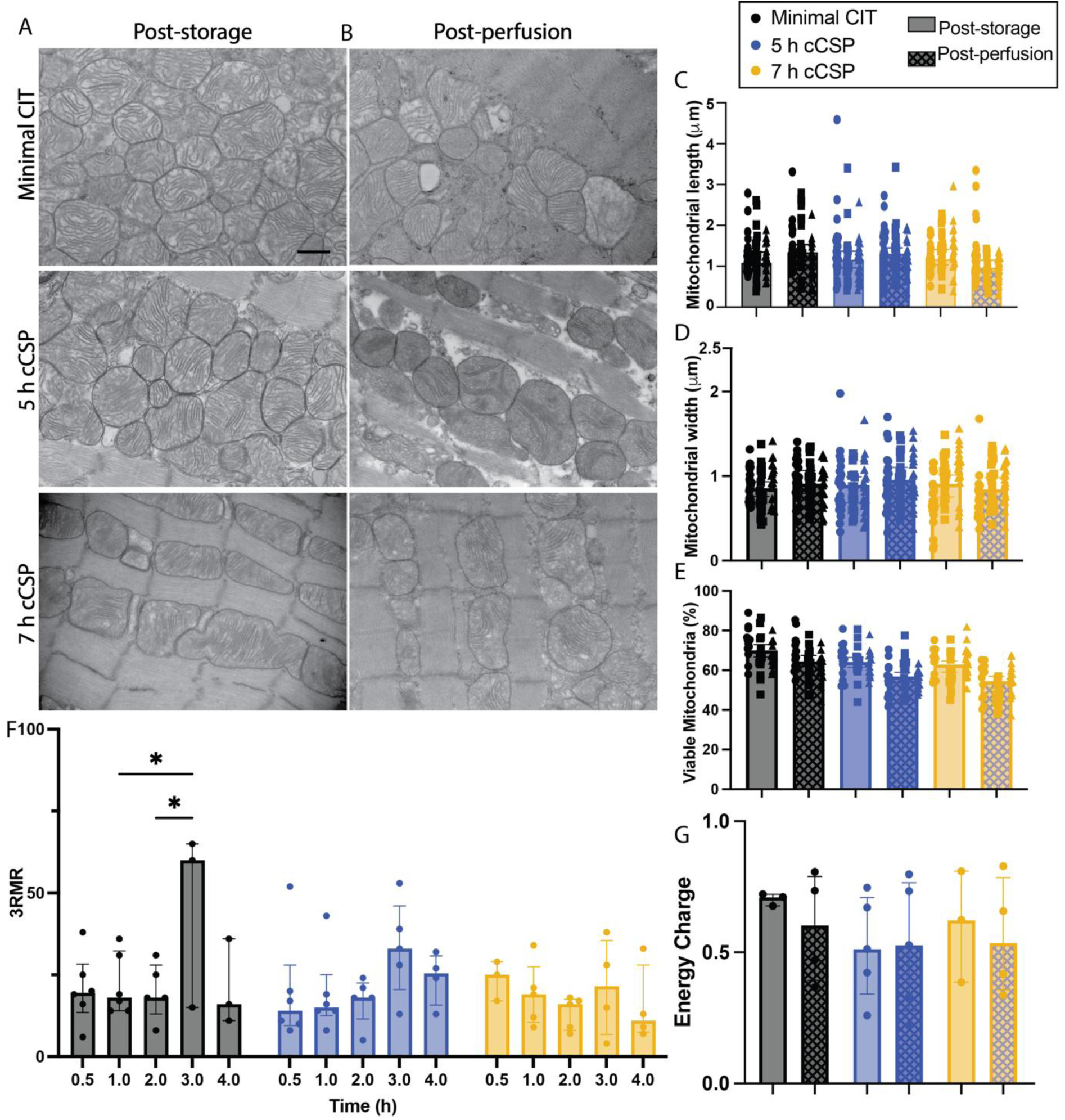
Mitochondrial ultrastructure, redox state and energy stores. Transmission electron microscopy (TEM) representative images of right ventricular tissue (A) post-storage and (B) post-perfusion. Scale bar 600 nm. Mitochondria (C) length and (D) width were quantified by measuring 20–25 mitochondria per TEM image of three biological replicates. Data points from each biological replicate are represented by different symbols. Solid columns indicate post-storage measurements, checkered columns indicate post-perfusion measurements. (E) Over time mitochondrial redox state with 3RMR determining the percent of reduced mitochondria. (F) Adenylated energy charge of right ventricular tissue post-storage (solid columns) and post-perfusion (checkered columns). Colum data presented as median ± interquartile range. Wilcoxon/Kruskal-Wallis Test with Dunn post-hoc analysis on non-parametric medians. Repeated measures two-way ANOVA and Tukey-Kramer HSD post-hoc analysis on standard least squares means. * = p <0.05.

### Controlled hypothermic preservation prior to NMP selectively attenuates the release of selective cytokines; NMP induces a robust inflammatory response in DCD hearts

Previous studies have demonstrated that recipients of DCD hearts preserved via NMP experience increased episodes of acute rejection and hospitalization post-transplant [43]. While the underlying mechanisms remain unclear, ischemia-reperfusion injury is known to trigger a potent inflammatory response that can influence graft tolerance and rejection [44]. Despite the storage time (i.e. longer cold ischemia exposure) cardiac grafts exposed to controlled hypothermia for 5 hours or 7 hours do not experience an additional increase in inflammatory response immediately after reperfusion (30 mins) when compared to cardiac grafts reperfused after minimal hypothermic storage (T0 Minimal CIT vs. T0 cCSP, Fig. 6). On the contrary, the accumulation of IL-1RA (Fig. 6A, p = 0.003) and IL-12 (Fig. 6F, p = 0.032) after 4 hours was significantly lower, while IL1β trended towards significance (Fig. 6C, p = 0.073) in the controlled hypothermia group, suggesting a blunted inflammatory response. Notably, both pro- (IL-1 receptor antagonist (IL-1RA), IL-10) and anti-inflammatory (IL1β, IL-6, IL-8, and IL-12) cytokines increased significantly over perfusion time in both groups (all p < 0.001), while IL-18 did not show a significant time-dependent increase. These results indicate a robust inflammatory response mediated by NMP.

**Figure 6:**
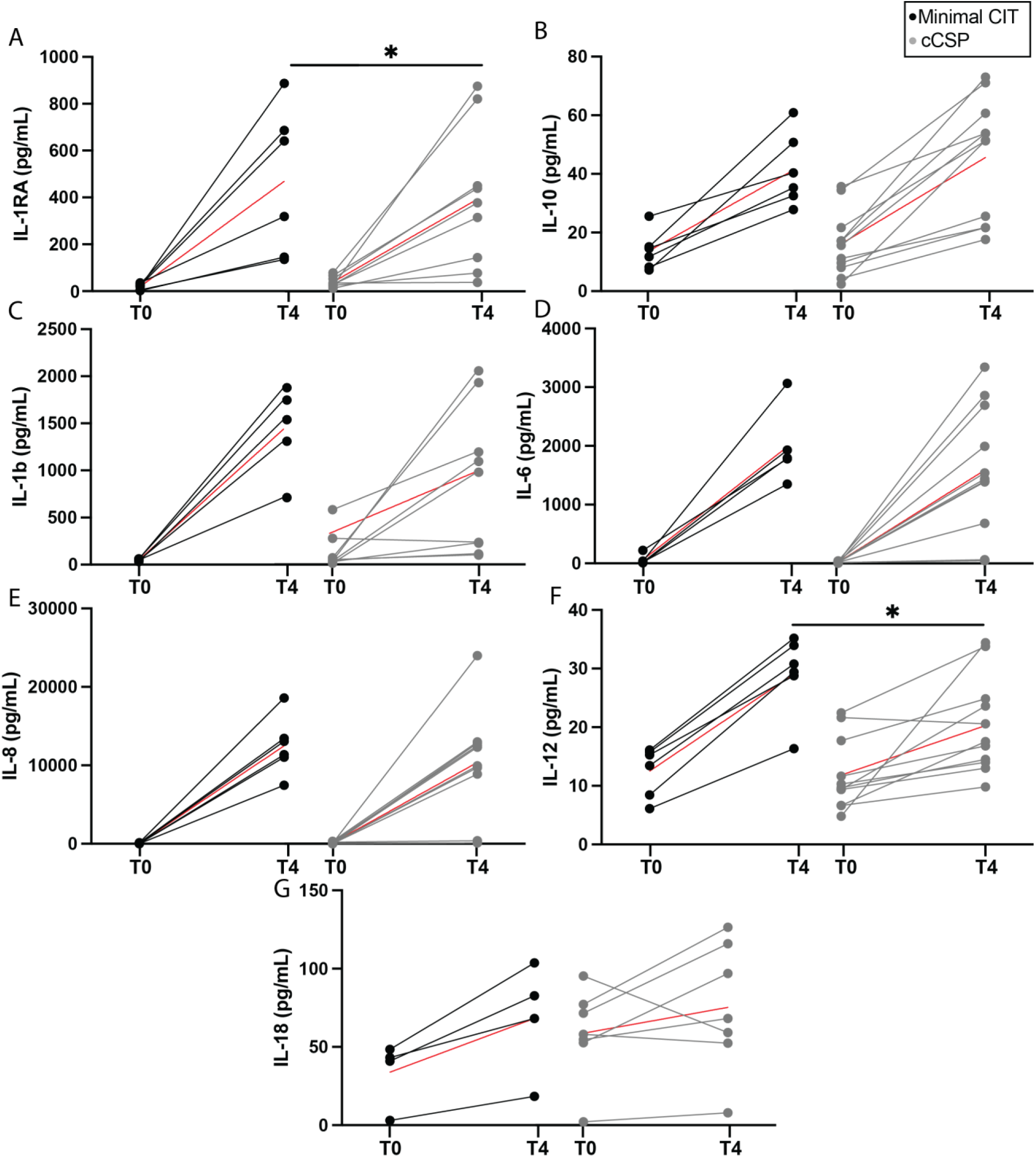
Pro and anti-inflammatory markers immediately after reperfusion (T0, 30 mins) and after 4 hours of NMP (T4). To increase statistical power, cytokine analysis was limited to two groups: Minimal CIT and controlled cold static preservation (cCSP). (A - B) In perfusate concentration of anti-inflammatory and (C – G) pro-inflammatory cytokines at 30 mins (T0) and 4 hours (T4) of perfusion time. Median is represented by red line. Data analyzed via difference in difference test. * = p <0.05.

### Controlled hypothermic preservation does not alter the myocardial proteome before or after NMP

Preservation strategies and prolonged ischemia are known to influence the epigenetic programming of cellular stress responses, which can manifest through transcriptomic and proteomic changes. While some effects may be apparent immediately after reperfusion— structurally, ultrastructurally, or functionally—others may impact graft performance later, particularly post-implantation. Proteomic analysis offers an early window into these molecular changes. Principal Component Analysis of the proteome profile of the right ventricle (RV) show overlap between non-stored grafts (Minimal CIT) and those stored via cCSP post-storage/pre-perfusion (Fig. 7A), and no significant differentially abundant proteins were found (Fig. 7D). Similarly, no differences in the proteome profiles were detected in either the right (Fig. 7B & 7E) or the left (Fig. 7C & 7F) ventricle between Minimal CIT grafts and cCSP grafts after 4 hours of NMP, with the exception of CYFIP related Rac1 interactor A (CYRIA), which is upregulated after NMP in the RV. Overall, these results indicate that controlled hypothermia does not significantly affect the proteome profile of DCD grafts.

**Figure 7:**
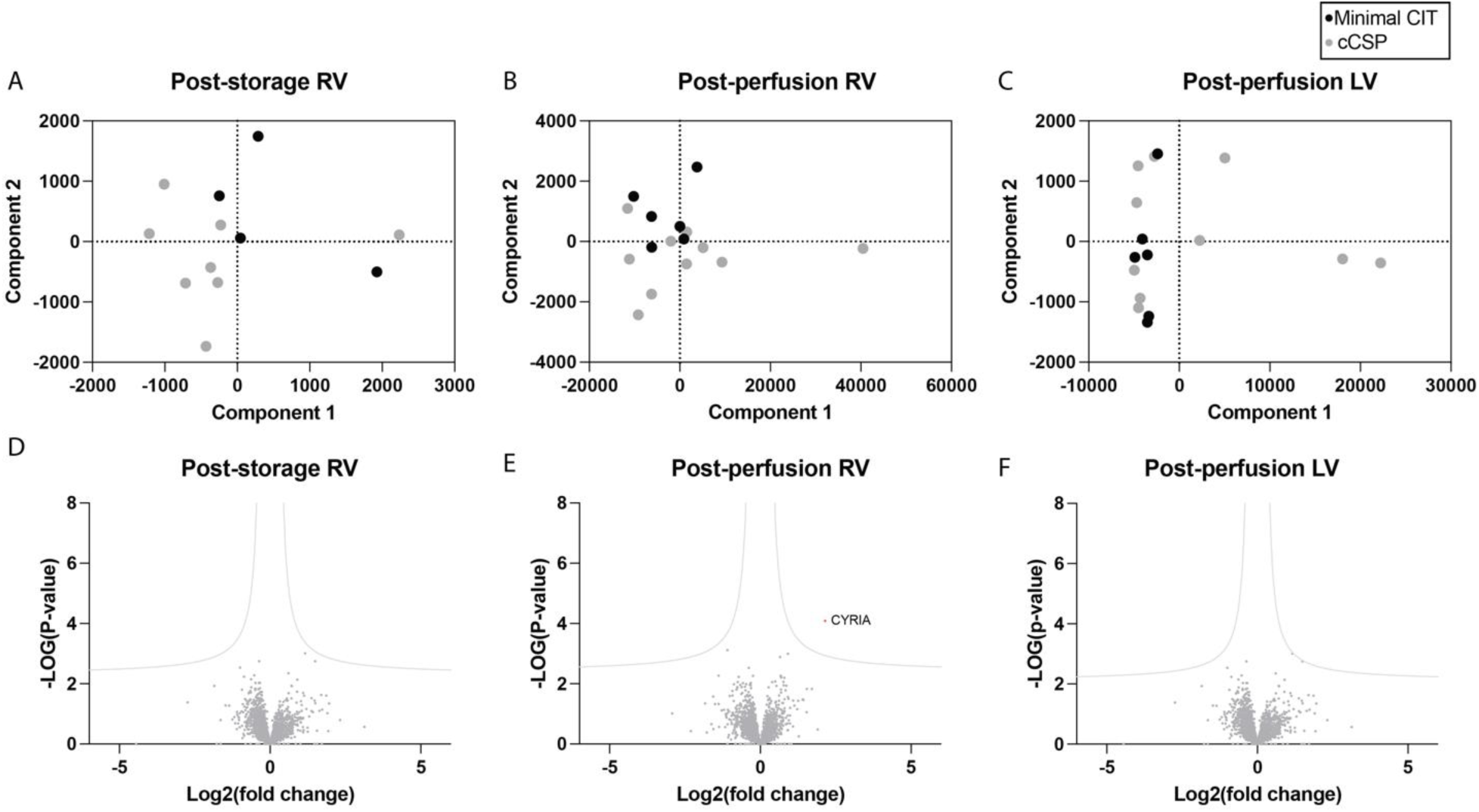
Effect of CIT on the myocardium proteome. To increase statistical power, proteomic analysis was limited to two groups: Minimal CIT and controlled cold static preservation (cCSP). Each dot on the PCA graphs represent a biological replicate. Among the post-storage cCSP samples, n = 6 were 5-h cCSP and n = 2 were 7-h cCSP. Among the post-perfusion cCSP samples, n = 5 were 5-h cCSP and n = 5 were 7-h cCSP. Principal Component Analysis (PCA, panels A-C) and Volcano Plots (panels D-F) were generated from the proteome profiles of post-storage RV and post-perfusion RV and LV.

Alternatively, NMP seems to cause a differential effect in the proteome irrespective of storage, with post-perfusion proteome profiles (open circles) of both Minimal CIT (Fig. 8A) and cCSP (Fig. 8B) visibly clustering in distinct groups than their respective post-storage profiles (filled circles). Interestingly, however, exposure to controlled hypothermia seems to alter the magnitude of the effect to the proteome, with Minimal CIT grafts differentially expressing only 51 proteins (Fig. 8C, Suppl Table 1, Suppl Fig. 1), whereas 324 proteins were differentially expressed in cCSP grafts (Fig. 8D, Suppl Table 2, Suppl Fig. 2). Of the 51 differentially expressed proteins in Minimal CIT grafts, 39 are also differentially expressed in cCSP grafts.

**Figure 8:**
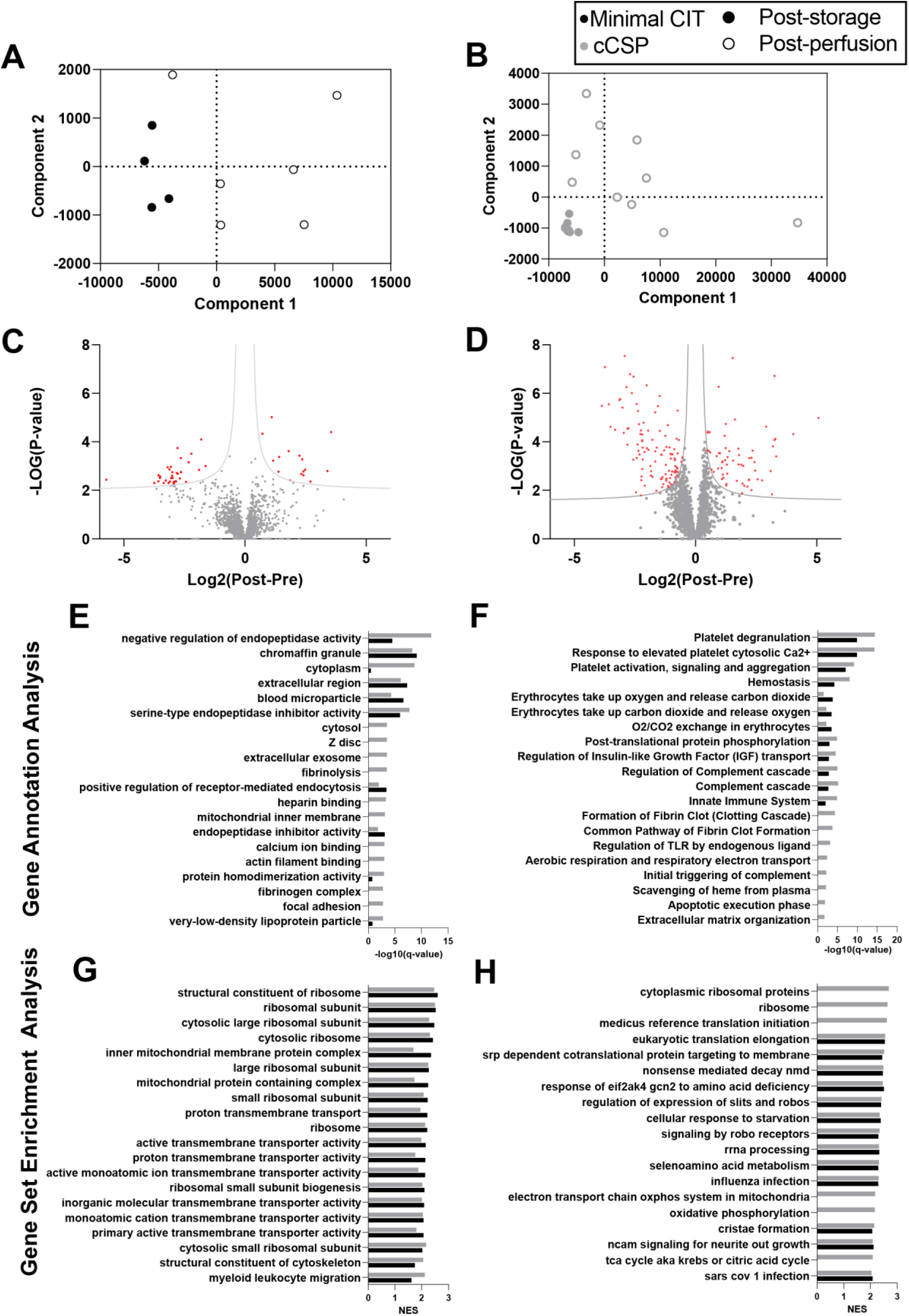
Effect of NMP on myocardium proteome profiles. To increase statistical power, proteomic analysis was limited to two groups: Minimal CIT and controlled cold static preservation (cCSP). Each dot on the PCA graphs represent a biological replicate. Among the post-storage cCSP samples, n = 6 were 5-h cCSP and n = 2 were 7-h cCSP. Among the post-perfusion cCSP samples, n = 5 were 5-h cCSP and n = 5 were 7-h cCSP. Principal Component Analysis (PCA, panels A-B) and Volcano Plots (panels C-D) were generated from the proteome profiles of RV post-storage and post-perfusion. The PCA graphs show that post-perfusion (open circles) proteome profiles cluster in distinct groups when compared to their respective post-storage (filled circles) profiles, indicating changes in proteome caused by NMP. Volcano plots revealed (C) 51 differentially expressed proteins before and after perfusion of Minimal CIT grafts, and (D) 324 differentially expressed proteins post-storage/pre-perfusion and post-perfusion of grafts stored in cCSP as determined by Student’s t-test and multiple adjustment by permutation based false discovery rate. The gray line represents the line of significance for a false discovery rate of 0.05 and a fold change cutoff s0 of 0.1.

Qualitative gene ontology analysis was performed on the 51 and 324 proteins whose abundance was affected by NMP in minimal CIT and cCSP grafts, respectively. In Minimal CIT grafts, NMP mostly affected proteins annotated with Gene Ontology (GO) terms related to circulating proteins (e.g., extracellular space/region, vitamin transport) erythrocytes (e.g., iron ion transport, oxygen binding), and platelets e.g., platelet degranulation, complement activation; Fig 8E, Suppl Table 3). Similarly, the proteins primarily affected by NMP is Minimal CIT grafts were annotated with Pathways terms related to red blood cell pathways (platelet degranulation, platelet activation, hemostasis, O_2_/CO_2_ uptake; Fig 8E, Suppl Table 4). For hearts stored in cCSP, NMP resulted in an over-representation of proteins annotated with GO terms and pathways related to plasma, erythrocytes and platelets (Fig 8F, Suppl Tables 5-6). However, NMP in cCSP hearts also affected proteins related to inflammation (innate immune system, Regulation of TLR by endogenous ligand), the mitochondria (mitochondrial inner membrane, aerobic respiration and respiratory electron transport), and myocyte cytoskeleton (Z disc, actin filament binding; Fig 8F, Suppl Tables 6-7).

GO and Pathways term enrichment analysis on low amount of differentially expressed proteins may lack statistical power. Therefore, a gene set enrichment analysis (GSEA) was performed, which considers ranked gene expression data and may provide more robust insights in this context, especially for the minimal CIT grafts. GSEA showed that NMP primarily affects proteins related to ribosomal activity, transporter activity and mitochondria in minimal CIT grafts (Fig 8G, Suppl Table 7). The top 15 canonical pathways affected by NMP in these hearts were related to translation, metabolism, inflammation, and the mitochondria (Fig 8G, Suppl Table 8). Interestingly, the GSEA analysis showed that GO terms and pathways affected by NMP in cCSP hearts were dominated by ribosomal function, transcription and translation (11 and 5 out of 15 most affected GO terms and canonical pathways, respectively; Fig 8H, Suppl Tables 9-10).

## Discussion

This study demonstrates that controlled hypothermic (6–8°C) preservation of DCD hearts prior to normothermic machine perfusion (NMP) maintains graft function, metabolic integrity, and structural preservation at levels comparable to the current clinical standard of immediate NMP following procurement. These findings challenge the prevailing assumption that any delay between DCD procurement and reperfusion is inherently harmful, and support a potential paradigm shift towards a simpler, more flexible, and potentially more scalable strategy for preservation and evaluation of donated DCD hearts.

A notable strength of controlled hypothermic storage is the logistical flexibility it introduces. Unlike the direct procurement and perfusion (DPP) approach, which requires immediate reperfusion and reliance on expensive, portable NMP devices, controlled cold storage allows for organ transport without a running perfusion system, significantly reducing cost, personnel demands, and time sensitivity. In addition, reanimation at the recipient center enables the use of more sophisticated ex vivo assessment platforms, including physiologic loading, to better evaluate graft viability prior to transplantation [29]. Furthermore, the combination of controlled hypothermic storage with subsequent NMP assessment resulted in an important extension of out-of-body time, which permitted an unprecedented 12 hours (1 hour instrumentation + 7 cCSP + 4 NMP) without any loss in metabolic fitness (Fig. 2) or function (Fig. 3). This unintended yet impactful outcome has the potential to address a second major hurdle in the field of organ transplantation: the persistent organ shortage crisis, by enabling broader geographic sharing, reducing discard rates, and increasing the overall pool of transplantable hearts.

In addition to its logistical advantages, controlled hypothermia also appears to induce a state of reduced protein synthesis even after reanimation and 4 hours of NMP. In terms of immune activation, controlled hypothermia may contribute to delayed immune recognition and lower acute rejection risk post-transplant, as an important downregulation in the protein expression of a wide array of immune-related genes was seen after NMP in both minimal CIT grafts and grafts stored in cCSP (Suppl Table 2, Supp. Fig. 2 and 4). However, a larger number of key components of the complement system saw their expression reduced in cCSP grafts after NMP compared to minimal CIT grafts, including, complement factor I, heparin cofactor II (SERPIND1), and prothrombin. Likewise, several acute-phase proteins were reduced after reperfusion in cCSP grafts but not in minimal CIT grafts, such as SERPINA1, fibrinogen chains FGA, FGB and FGG, and haptoglobin, all of which play significant roles in immune activation and inflammatory responses [45, 46]. This suppression of immunogenicity by hypothermic preservation could potentially minimize the risk of immune-mediated injury, as its inhibition has been associated with reduced likelihood of graft rejection after transplantation in other organs [47].

In parallel, hypothermic preservation also led to the downregulation of several transcriptional regulators involved in driving inflammatory gene expression. Specifically, proteins such as general thyroid hormone receptor-associated protein 3, nuclear factor 1, and KH-type splicing regulatory protein which collectively influence the activation and stability of pro-inflammatory transcripts including cytokines and complement factors, were significantly suppressed in cCSP-preserved hearts. The attenuation of these regulators may contribute to the observed decline in inflammatory protein expression, reinforcing the concept of hypothermia-induced immune quiescence. The effects of reduction of these proteins can be seen in the significant reduction of the interleukin-mediated inflammatory response associated with ischemia–reperfusion injury [48], as evidenced by the decreased accumulation of select pro-inflammatory mediators such as IL-1RA and IL-12 in grafts preserved under controlled hypothermia (Fig. 6). In conjunction, these gene suppressions can likely create a low-immunogenic, low-inflammatory profile, which may contribute to delayed immune recognition and lower acute rejection risk post-transplant.

While the downregulation of inflammatory and immune pathways during hypothermic storage likely contributes to reduced graft immunogenicity, the sustained suppression of genes critical for cardiac function may hinder post-transplant recovery. Persistent downregulation of key contractile and structural proteins, such as myosin-6, myosin light chain kinase, and myosin light chain 12A, may impair the reestablishment of myocardial contractility, limiting the heart’s ability to generate sufficient force under post-reperfusion stress [49, 50]. However, GSEA analysis shows that the gene set structural constituent of cytoskeleton is enriched in cCSP hearts after NMP, with cytoskeletal-associated proteins driving this enrichment (e.g., desmin and tubulin beta 3, desmoplakin, which are not expressed by circulating cells). Similarly, reduced expression of proteins involved in cardioprotective signaling, cellular repair, and mitochondrial dynamics (mitogen-activated protein kinase, catenin beta-1, and dynamin-1-like protein), could delay adaptive responses required for recovery from ischemia–reperfusion injury. On the other hand, GSEA analysis revealed that the gene set monoatomic cation transmembrane transporter activity was enriched in cCSP grafts, driven by SURF1, a protein involved in cytochrome C oxidase assembly (Complex IV of the electron transport chain, ETC), mitochondrially encoded cytochrome b, a core protein of complex III of the ETC and a gene encoded by mitochondrial DNA, NADH Ubiquinone oxidoreductase subunit A8, part of Complex I of the ETC. While immune quiescence is a desired effect of hypothermic storage, a timely reactivation of essential contractile, metabolic, and repair pathways is likely critical for full functional reanimation and long-term graft viability.

As lesser-known side effects of cold storage, such as protein down regulation, are increasingly recognized, greater attention must be given to the temperatures at which organs are preserved. Slightly higher storage temperatures may be more beneficial than conventional static cold storage (0–4°C), offering effective preservation while potentially allowing for the reversible suspension of organ functions. Excessive cooling, such as that seen during conventional static cold storage, has been shown to exacerbate inflammatory responses, increase structural damage upon reperfusion, and contribute to adverse post-transplant events [40, 51, 52]. Although a downregulation of proteins key to myocardial function is observed in this study, it is interesting to note that gene sets related to protein synthesis are enriched in cCSP grafts after NMP, including translation, RNA processes, and ribosomal functions (Suppl Table 9-10). In fact, 11 out of the top 15 GO terms were related to protein synthesis in cCSP grafts, while only 6 were found in minimal CIT grafts. This suggests that there is more protein synthesis in cCSP grafts when compared to minimal CIT grafts. This underscores the importance of precise temperature control which may provide a more favorable balance between metabolic suppression and organ integrity.

Although, not unexpected, it is equally important to note the heighten inflammatory response NMP seem to modulate in both Minimal CIT and cCSP hearts with both pro- and anti-inflammatory markers exhibiting up to a thousand-fold increase at the end of 4 hours of perfusion when compared to initial perfusion time (Fig. 6), consistent with findings reported in other studies [31, 53]. Similarly, NMP appears to cause an important deterioration in the structural integrity of ventricular tissue, as observed in H&E images (Fig. 4), not evident in the tissue post-preservation/pre-perfusion. While cytoplasmic vacuolation can be possible attributed to ischemia [54], interstitial edema and vascular congestion are a likely consequence of the inflammatory milieu generated during perfusion and the non-physiological conditions of normothermic machine perfusion [55].

Another challenging aspect of the experimental setup used in this study was replicating circulatory death in the porcine model. Swine hearts are known to exhibit a heightened sensitivity to ischemia compared to human hearts, which presents a unique limitation in the direct translation of human protocols into swine experimental models [56]. This increased vulnerability is largely attributed to the pig myocardium’s higher metabolic rate and reduced ischemic tolerance, leading to a faster onset of irreversible myocardial damage [57, 58]. As a result, even brief periods of warm ischemia can significantly impair cardiac function in pigs [59, 60]. Therefore, while the onset of functional warm ischemic time (fWIT) in humans is typically defined by a sustained systolic blood pressure of less than 50 mmHg [61], systemic oxygen saturation (SpO_2_) is often disregarded due to the unreliability of measurements in the absence of pulsatile flow. However, SpO_2_ at the onset of fWIT may be critical to the success of the model and should not be overlooked in porcine studies, as SpO_2_ values below approximately 70% at the initiation of the 19-minute fWIT have been associated with non-reanimatable grafts (Supp. Table 11)

### Limitations

This study has several limitations. First, while the ex vivo platform utilized allows for detailed physiologic assessment, only the left ventricle was loaded. As a result, functional evaluation of the right ventricle (RV) was limited. This represents a significant gap, particularly in the context of DCD transplantation, where the RV is known to be more vulnerable to ischemia-reperfusion injury and often exhibits a higher incidence of early post-transplant failure. Second, although this study focused on early reperfusion and assessment endpoints, post-transplant function or survival was not evaluated. Therefore, a full transplant, longitudinal experimental setup would provide more comprehensive insight into biventricular performance, immune activation, and longer-term outcomes, crucial to confirming these findings and assessing the true clinical utility of this strategy.

Lastly, the results for GO and GSEA analyses must be interpreted carefully due to the lower n value for Minimal CIT hearts vs. cCSP. Furthermore, the addition of blood proteome to the myocardium proteome during NMP, makes it difficult to determine if the identified differentially abundant proteins are due to the addition of blood or to the myocardium’s response to NMP. In fact, the analyses of GO and pathway annotations (Suppl Tables 3 to 6) shows that GO terms related to blood are dominant (e.g., extracellular space, blood microparticle, oxygen binding, platelet degranulation, hemostasis). However, as RBCs are devoid of intracellular organelles, the enrichment of ribosome and mitochondria proteins are likely due to changes in the myocardium proteome. This can also be inferred when looking at the GSEA analysis, where terms unrelated to blood are dominant, including protein synthesis (ribosome, eukaryotic translation elongation), transporter activity, mitochondrial function (cristae formation, mitochondrial protein import), and inflammatory response (myeloid leukocyte migration, IL4 and IL13 signaling). Since GSEA offers a more comprehensive picture of changes in the proteome, it is important to consider its results alongside annotation analyses to capture both subtle and pronounced biological shifts.

## Conclusion

The results in this study demonstrate that DCD hearts preserved under moderate hypothermia for up to 7 hours retain functional, structural, and metabolic viability equivalent to those reperfused immediately. This approach offers a logistically simpler and more cost-effective alternative to current DPP practices, with the potential to reduce inflammation and preserve mitochondrial integrity. These findings support a reevaluation of existing DCD heart preservation protocols and lay the groundwork for further translational studies aimed at optimizing heart transplant logistics and outcomes.

## Data Availability

Data will be available upon request

## Acknowledgements

This work was supported by generous funding to S.N.T. from the US National Institutes of Health (K99/R00 HL1431149; R01HL157803). We also gratefully acknowledge funding from the US National Institute of Health (R01DK134590; R24OD034189), National Science Foundation (EEC 1941543), American Heart Association (18CDA34110049), Harvard Medical School Eleanor and Miles Shore Fellowship, Polsky Family Foundation, the Claflin Distinguished Scholar Award on behalf of the MGH Executive Committee on Research, and Shriners Children’s Boston (Grant #BOS-85115). We also thankfully acknowledge support for S.A.R Polsky Family Foundation. The authors would also like to thank the Center for Comparative Medicine and the Knight Surgical Research Lab at Massachusetts General Hospital for animal management. We thank the Mass Spectroscopy, Genomics and Proteomics, and Morphology facilities at Shriners Children’s Boston. Electron microscopy was performed in the Microscopy Core of the Program in Membrane Biology, which is partially supported by an Inflammatory Bowel Disease Grant DK043351 and a Boston Area Diabetes and Endocrinology Research Center (BADERC) Award DK135043.

---

**Suppl Figure 1:**
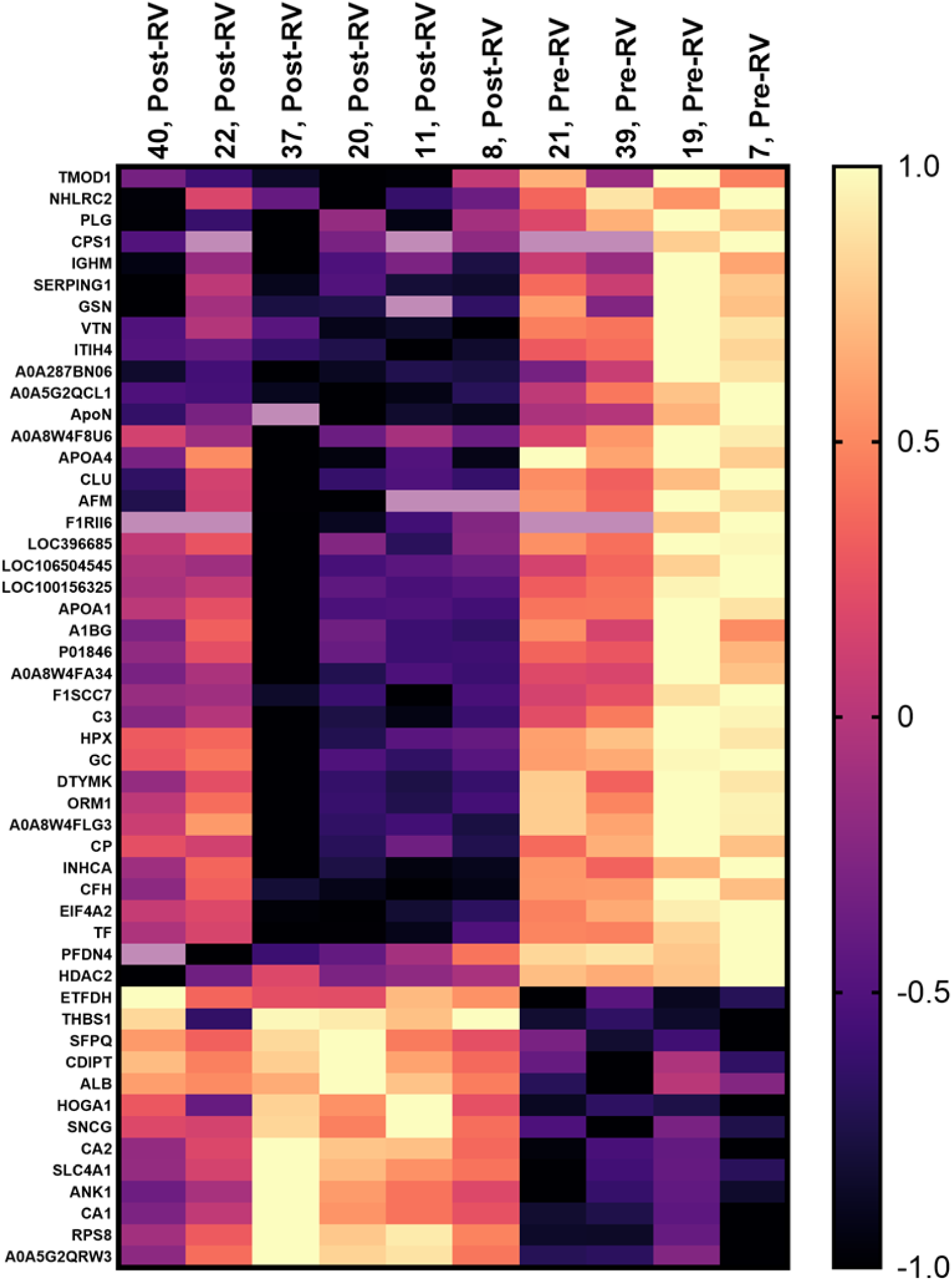
Heatmap of the 51 differentially abundant proteins in the RV of minimal CIT grafts post NMP compared to pre-NMP, as determined by Student’s t-test and multiple adjustment by permutation based false discovery rate.

**Suppl Figure 2:**
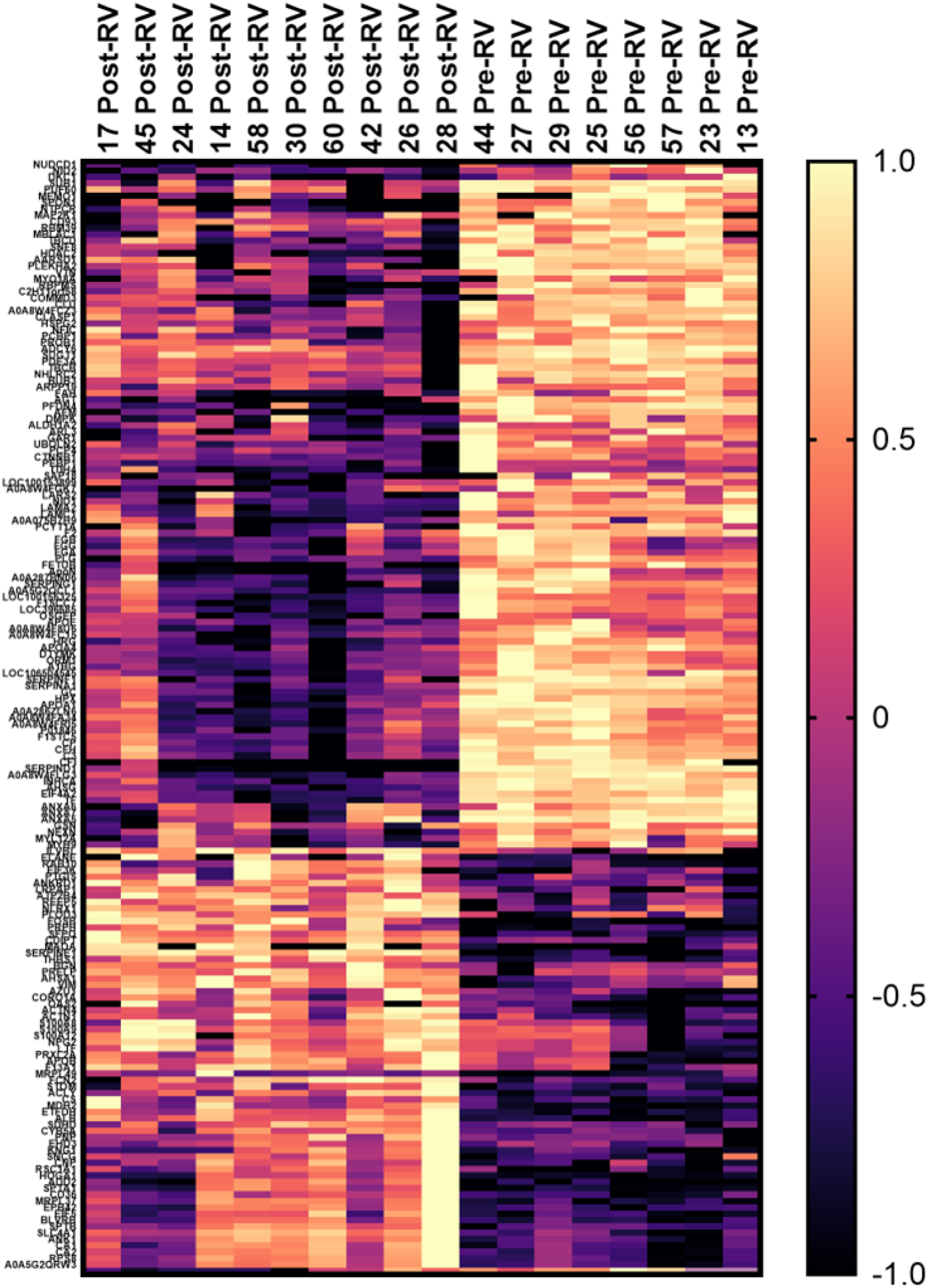
Heatmap of the 324 differentially abundant proteins in the RV of cCSP grafts post NMP compared to pre-NMP, as determined by Student’s t-test and multiple adjustment by permutation based false discovery rate.

**Suppl Figure 3:**
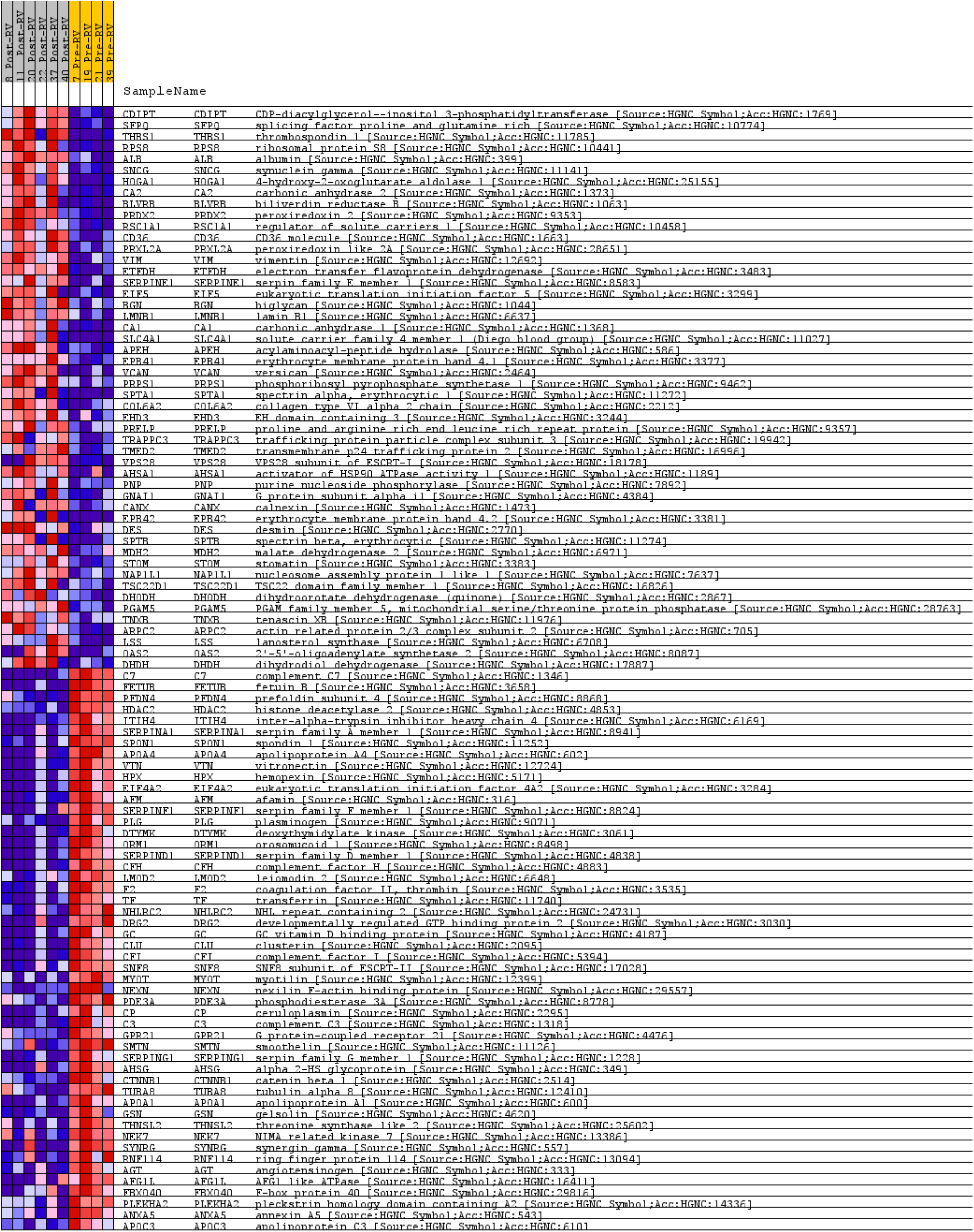
Heatmap generated after GSEA analysis of Minimal CIT RV before (pre-RV) and after (post-RV) NMP.

**Suppl Figure 4:**
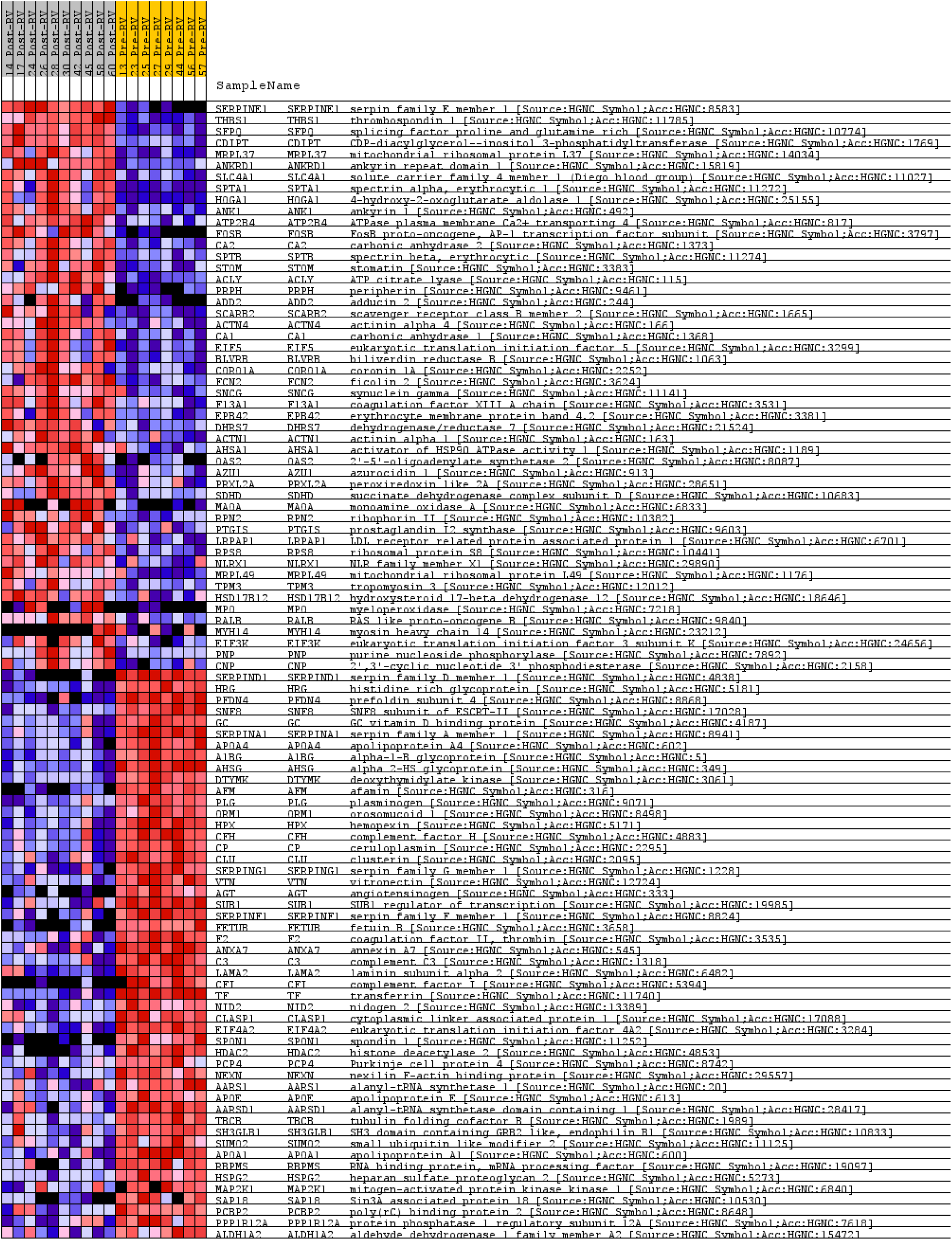
heatmap generated after GSEA analysis of cCSP RV before (pre-RV) and after (post-RV) NMP

